# Temporal Summation of Subthreshold Stimuli in Human Motor Axons: Implications for Intraoperative Neuromonitoring

**DOI:** 10.1101/2025.08.13.25333601

**Authors:** A Naidoo, KE Jones

**Author notes:** Corresponding author: Kelvin E Jones University of Alberta Faculty of Kinesiology, Sport, & Recreation Van Vliet Center 4-220 116 St and 85 Ave Edmonton, Alberta T6G 2R3.

## Abstract

**Objectives:** To examine how stimulus amplitude and width influence subthreshold superexcitability of peripheral axons and to provide evidence-based recommendations for minimizing inadvertent compound muscle action potential (CMAP) generation during intraoperative corticobulbar monitoring.

**Methods:** Fifteen healthy participants received median nerve stimulation under nine conditions combining three amplitudes (80%, 85%, and 90% of threshold) and three pulse widths (0.1 ms, 0.5 ms, 1.0 ms). Trains of 1–7 subthreshold pulses (2 ms interpulse interval) were delivered 10 times per condition. CMAPs were recorded from the abductor pollicis brevis (APB), and the probability of a response exceeding 100 µV (baseline-to-negative-peak) was calculated. Persistent sodium current was estimated using the latent addition test (LAh).

**Results:** Higher pulse amplitudes and wider pulse widths significantly increased CMAP probability, with a significant interaction (F(4,56) = 4.853, p = .002, partial η² = .257). All pairwise comparisons were significant (p ≤ .023). When controlling for rheobase, LAh was positively correlated with response probability (r_partial_(12) = .539, p = .047).

**Conclusions & Significance:** Subthreshold trains activate motor axons in a predictable manner depending on amplitude, width, and train length. These findings challenge current IONM assumptions and highlight the need for threshold-referenced, standardized protocols.

## 1. Introduction/Background

Intraoperative neuromonitoring (IONM) of cranial nerves, particularly the facial nerve, is widely recognized as important for preserving neural function and reducing the risk of postoperative neurological deficits during brainstem and skull base surgeries (Chang et al., 1999; Isaacson et al., 2005; Neuloh et al., 2008). A standard approach is transcranial electrical stimulation (TES) delivered over lateral scalp positions to activate the precentral gyrus (Goto et al., 2010). However, TES can inadvertently excite peripheral portions of cranial motor nerves, leading to the generation of compound muscle action potentials (CMAPs) in target muscles even when the corticobulbar pathway is disrupted. This phenomenon risks misinterpretation of a CMAP as a true corticobulbar motor-evoked potential (MEP), potentially resulting in a false-negative assessment of neural integrity (Akagami et al., 2005; Dong et al., 2005).

The potential for peripheral activation during corticobulbar MEP monitoring was recognized early in the development of IONM protocols (Dong et al., 2005; Deletis et al., 2016). To distinguish central from peripheral responses, a verification step was introduced: if a train of stimuli elicits a muscle response but a single stimulus of the same intensity does not, the response is attributed to transsynaptic corticobulbar activation. This interpretation assumes that axonal excitability is fixed; specifically, that subthreshold stimuli have no cumulative effect on the axon. Under this assumption, if a single pulse fails to activate the axon, then a multipulse train at the same subthreshold intensity should also fail to evoke a response. However, this idea was challenged by work showing that repeated subthreshold stimulation can summate to directly activate motor axons (Téllez et al., 2016; Urriza et al., 2016; Urriza, 2020). In these studies, while a single subthreshold pulse never evoked a CMAP, trains of identical subthreshold pulses consistently did.

The physiological basis of this phenomenon, termed subthreshold superexcitability, was characterized by Bostock and colleagues in single human motor axons (Bostock et al., 2005). They showed that subthreshold electrical stimuli can transiently lower the threshold for action potential initiation. This effect includes a passive electrotonic component, proportional to the charge delivered by the stimulus, and an active component involving voltage-dependent channels. One potential contributor to the active component is the persistent sodium current, a slowly inactivating conductance that activates near the resting membrane potential (Crill, 1996; Baker and Bostock, 1997; Goldin, 2003). When activated by subthreshold stimuli, these channels can produce a sustained depolarizing drive, further reducing the threshold and increasing the likelihood of axonal firing during subsequent stimulus pulses.

Building on the foundational work of Urriza et al., who showed that subthreshold trains (90% of threshold, 0.5 ms pulse width) could evoke CMAPs in the abductor pollicis brevis, we sought to extend these findings by systematically varying both subthreshold pulse amplitude and width. Unlike prior studies, which focused on representative EMG traces, we quantified response probability across trials and participants to assess the likelihood of CMAP generation from subthreshold stimulation. Our objectives were to determine how stimulus parameters affect axonal excitability, to quantify individual susceptibility to subthreshold superexcitability, and to explore whether this susceptibility correlates with persistent sodium currents. To that end, we included a latent addition (LAh) test to estimate the contribution of persistent sodium conductance to the observed excitability patterns.

## 2. Methods

### 2.1 Participants

Fifteen healthy participants (9 male and 6 female), aged 21-28 years, were recruited through convenience sampling. All participants provided written informed consent. Exclusion criteria were adopted from the MScan Multicenter protocol (Sørensen et al., 2023). Prior to testing, participants completed a pre-screening to assess use of medications associated with peripheral neuropathy and to rule out the following: history of central nervous system disease or injury, upper limb injuries resulting in numbness or weakness, previous chemotherapy, diagnosis of diabetes mellitus, self-reported alcohol intake exceeding 14 units per week, other causes of polyneuropathy, known nerve entrapment syndromes, cognitive impairment, or pregnancy. All procedures were performed in compliance with relevant laws and institutional guidance, and the study was approved by the Research Ethics Board of the University of Alberta (Protocol #: Pro00142797).

### 2.2 Surface Electrode Placement

Three types of electrodes were used: stimulation (3M Red Dot, 3M-2560), recording (Ambu® BlueSensor NF), and ground (Ambu® Neuroline Ground). Participants were positioned in a reclined examination chair with their feet elevated. The median nerve was mapped for stimulation proximal to the wrist to identify the optimal site for the stimulating cathode.

Skin preparation at both stimulation and recording sites included application of Nuprep skin gel, 70% isopropyl alcohol, and light abrasion using 3M Red Dot Trace Prep sandpaper tape to maintain impedance at the skin-electrode interface below 10 kΩ. Following preparation, non-polarizable silver/silver chloride electrodes were placed over the mapped stimulation site; the return (anode) was placed 10 cm proximally.

Surface EMG was recorded from the abductor pollicis brevis (APB) muscle. The active recording electrode (E1) was positioned over the mid-belly of the APB and optimized to yield the largest, fastest-rising CMAP. The reference electrode (E2) was placed over the metacarpophalangeal joint, and the ground (E0) was positioned on the dorsum of the hand.

To maintain consistent nerve temperature, the forearm was wrapped with Norm-O-Temp water-heating blankets (Gentherm Medical, USA) set to 37 °C. Skin temperature was continuously monitored using a Digitimer D501 Isolated Temperature Monitor.

Stimulation was delivered via a Digitimer DS5 constant-current stimulator, controlled by QTracS software. Square-wave current pulses were applied to the median nerve. EMG signals were amplified (Digitimer D440-2; gain 300; bandpass 3–3000 Hz), mains noise suppressed using a HumBug 50/60 Hz noise eliminator (Quest Scientific), digitized (National Instruments USB-6251 BNC), and analyzed using QTracS.

### 2.3 Urriza Protocol

The “Urriza” protocol was implemented using QTracW software. Each participant completed a 3x3 repeated-measures factorial design, comprising nine unique conditions presented in randomized order. Independent variables were pulse amplitude (80%, 85%, and 90% of threshold) and pulse width (0.1, 0.5, and 1.0 ms). Threshold was determined separately for each pulse width and defined as the minimum stimulus intensity that evoked a CMAP with a baseline-to-negative peak amplitude ≥100 µV in 8 of 10 consecutive trials. Subthreshold amplitudes were then set to 80%, 85%, or 90% of that individual threshold.

For each condition, the subthreshold stimulus was delivered as trains of 1-7 pulses, each separated by a 2 ms interpulse interval (equivalent to 500 pulses per second). Each train was repeated 10 times with a 1 second interval between trains, and the number of CMAP responses ≥100 µV (baseline-to-negative-peak) out of ten was recorded. This criterion, ≥100 µV consistent with Urriza et al. (2016), provides an unambiguous CMAP well above background noise yet small enough to avoid excessive movement during intraoperative procedures. This full sequence was repeated three times per condition, with a 2 minute rest between cycles and approximately 5 minutes between each of the nine conditions. The entire protocol lasted about 90 minutes per participant. Figure 1 illustrates the workflow, timing and repetition structure. Response probability for each train length was calculated as the mean across the three repetitions, and a mean response probability for each participant was calculated as the average across all train lengths for each of the nine conditions (illustrated in Fig. 2).

**Fig 1.**
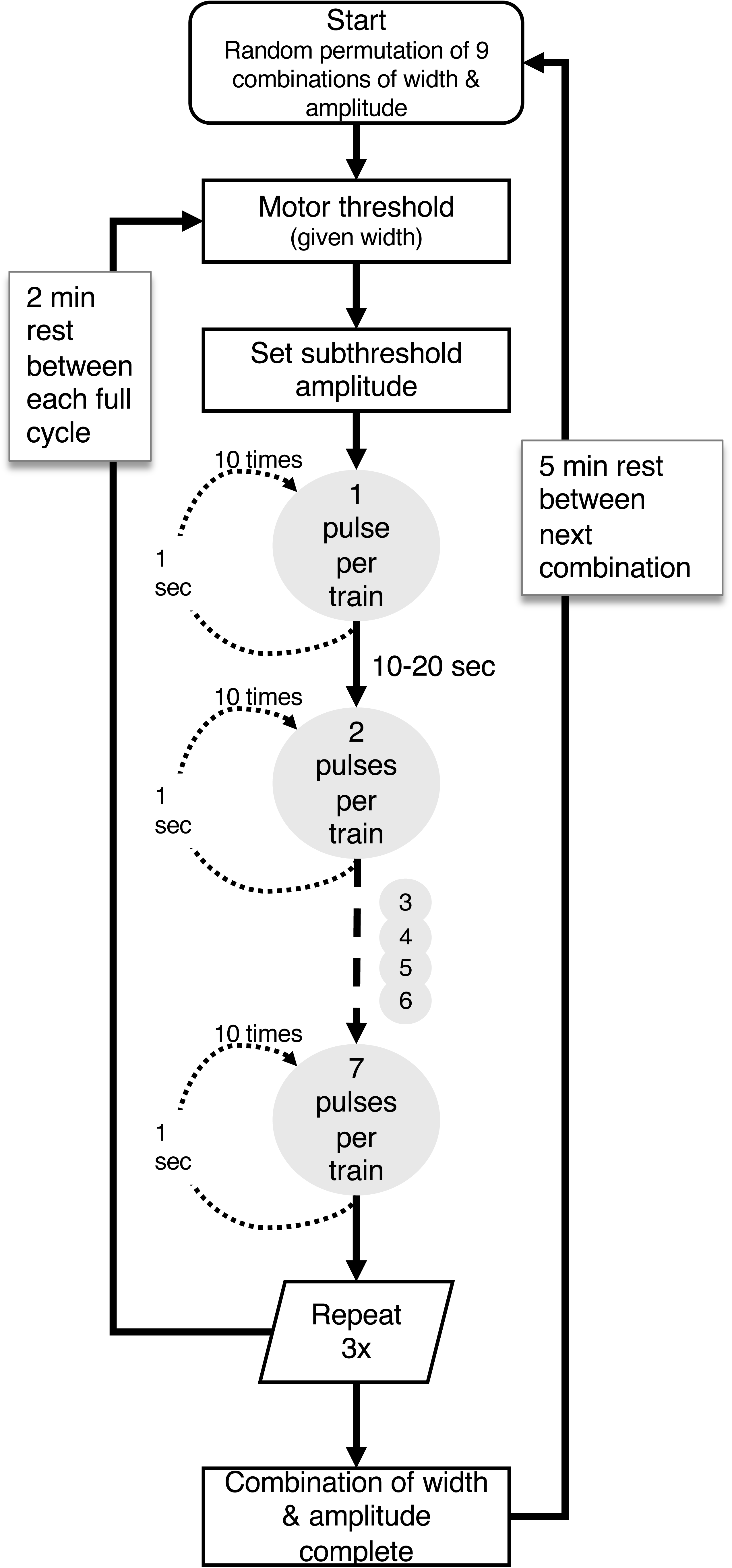
Flowchart of the electrical stimulation protocol. Each participant completed nine randomized conditions combining three pulse widths (1.0, 0.5, 0.1 ms) and three subthreshold amplitudes (90%, 85%, 80% of threshold). For each condition, the threshold amplitude for a given pulse width was defined as the current eliciting a CMAP (∼100 uV) in 8 of 10 stimuli. Subthrehold amplitude was then set to the designated percentage of this threshold. Bursts of 1–7 pulses (2 ms interpulse interval) were delivered 10 times, with 1 s between bursts and 10-20 seconds between burst lengths. The full burst sequence was repeated three times per width-amplitude combination, and the mean response probability for each burst length was calculated across the three cycles. This process was repeated for all nine width-amplitude combinations.

**Fig 2.**
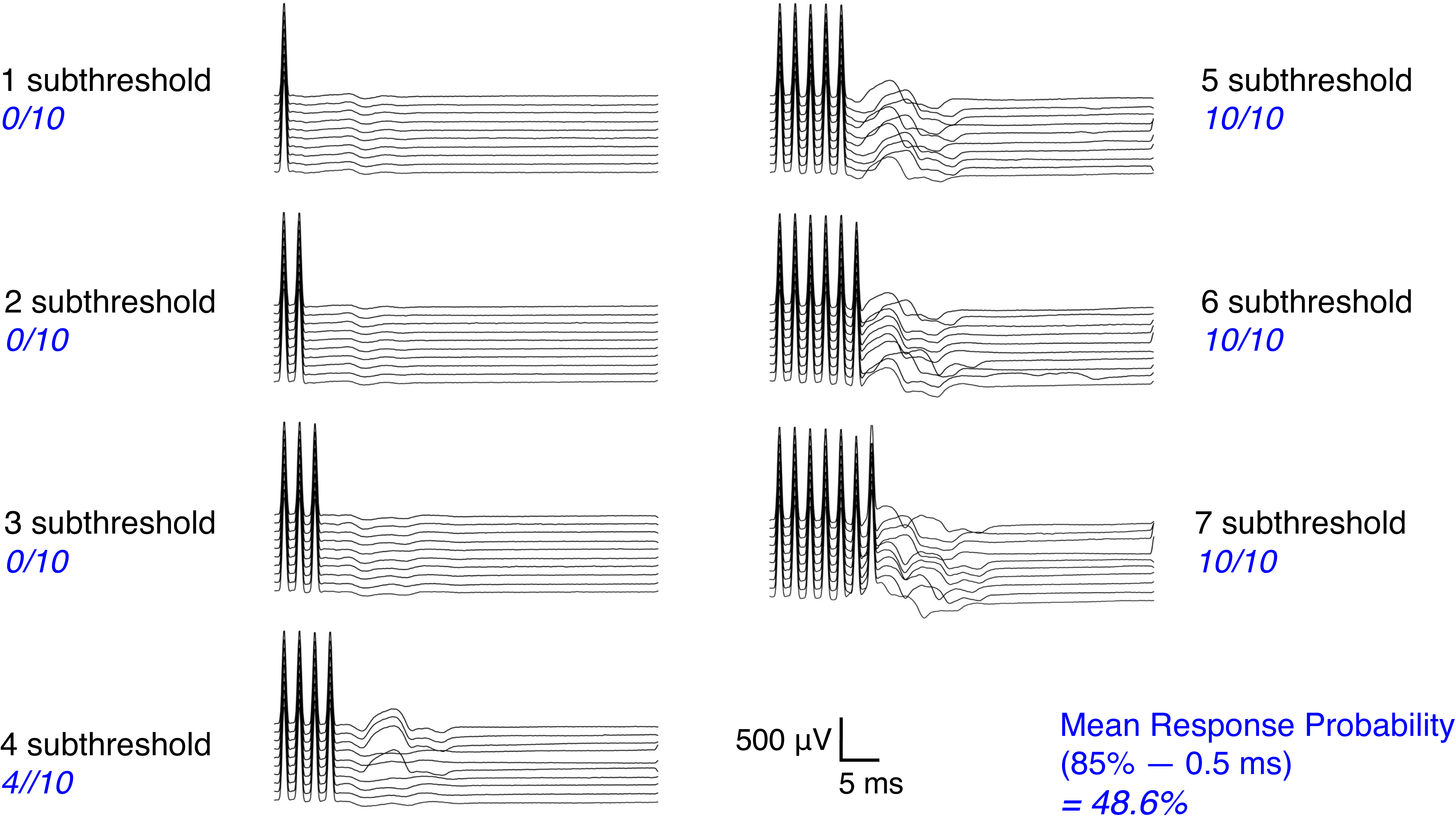
Example CMAP recordings from a single participant (APB muscle) showing responses to a single subthreshold pulse and to trains of 2-7 identical subthreshold pulses (0.5 ms pulse width, 85% of threshold intensity, 2 ms interpulse interval). The proportion of trials meeting the CMAP criterion (≥ 100 µV, regardless of latency) was used to calculate response probability. This illustrates how repeated subthreshold pulses can summate to trigger peripheral activation despite the absence of a single-pulse response.

### 2.4 Latent Addition Protocol

We used latent addition to estimate persistent sodium current because sustained inward current provides a biophysical mechanism for temporal summation between closely spaced subthreshold pulses (Bostock and Rothwell, 1997). In brief, the protocol measures the interaction between two 60 μs stimuli: a hyperpolarizing conditioning pulse followed by a test pulse, delivered at progressively increasing interstimulus delays. The conditioning pulse was set to 90% of the control threshold, defined as the current required to elicit a CMAP of 40% of maximum amplitude. At each delay, the test pulse intensity was adjusted to maintain the same 40% CMAP amplitude, and the resulting change in the test pulse threshold relative to the control, reflects the effect of the conditioning pulse. Threshold changes were expressed as a percentage of the control threshold and plotted on a semi-logarithmic scale as a function of interstimulus delay. The resulting time course was fitted with a biexponential function representing a fast passive component (τ_1_) and a slower active component (τ_2_) that has been linked to persistent sodium conductance.

The primary outcome of interest was the threshold reduction at a 0.2 ms delay (LAh), a time point at which the passive component is nearly extinguished, allowing selective quantification of the active component. LAh values were calculated for each participant to assess the magnitude of persistent sodium current.

### 2.5 Statistical Analysis

Statistical analyses were performed using SPSS (version 29). A two-way repeated measures ANOVA was performed to evaluate the effects of pulse amplitude (80%, 85%, and 90% of threshold) and pulse width (0.1 ms, 0.5 ms, and 1.0 ms) on the mean probability of response to subthreshold stimulus trains. For each participant, response probability was first averaged across the three cycles for each train length (1-7 stimuli), and then a condition-specific mean was calculated by averaging across train lengths for each width-amplitude combination. These condition-specific means were used in the repeated-measures ANOVA. When data were pooled across participants to illustrate group-level response curves, the median response probability was used for each train length to reduce the influence of outliers and to better represent the monotonic increase and saturation of response probability with additional pulses per train.

Assumptions for parametric statistical testing were evaluated prior to analysis. One outlier was identified among 135 data points (based on studentized residuals) but was retained as it did not affect the overall results. Shapiro-Wilk tests indicated one violation of normality (p < .05), but no data transformation was applied, given the robustness of repeated measures ANOVA to non-normality in within-subjects designs. Mauchly’s test confirmed sphericity for the interaction term. Significant interactions were followed up using simple main effects analysis with pairwise comparisons adjusted using the Bonferroni correction.

To characterize the sensitivity of motor axons to subthreshold trains of stimuli, we defined a subthreshold susceptibility index (SSI) as the number of pulses in a train required to elicit a CMAP response with a 5% probability. This 5% criterion was chosen to reflect an acceptable threshold for false-negative responses during intraoperative neuromonitoring, and was chosen based on statistical precedence. For each stimulation condition, the group median response probability was plotted against the number of pulses in the train. These data were fit to a three-parameter asymmetric sigmoid function:

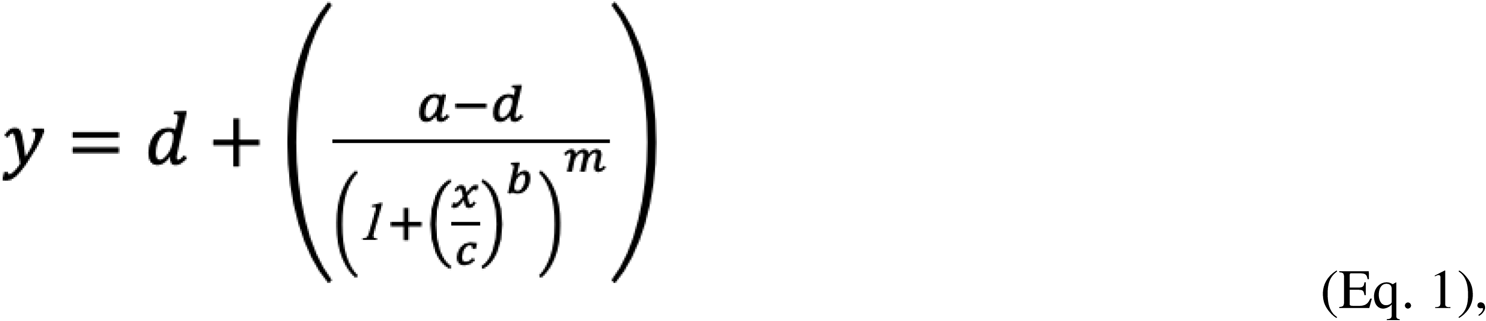

where *y* is the response probability, *x* is the number of stimuli, and constants were set to *a* = 0, and *d* = 100, reflecting a response range from 0 - 100%. Parameters *b*, *c*, and *m* were optimized by minimizing the sum of squared residuals using the GRG Nonlinear method (Lasdon et al., 1978). SSI was defined as the number of stimuli required to elicit a 5% response probability and was calculated by solving Eq. 1 for *x* when *y* = 5:

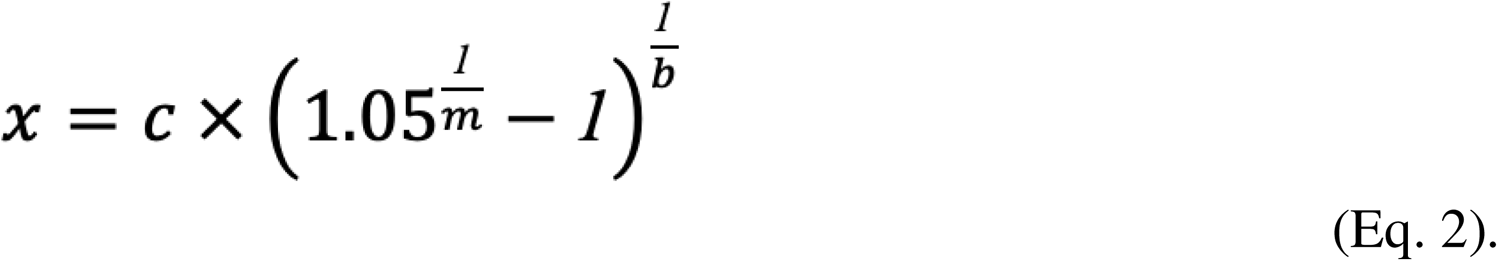

To examine whether persistent sodium currents contribute to inter-individual variability in subthreshold superexcitability, Pearson’s correlation was used to assess the relationship between each participant’s mean probability of response and their LAh value. Because rheobase provides a general index of baseline excitability and may influence response probability independent of LAh, a partial correlation controlling for rheobase as a covariate was also performed. This adjustment was intended solely to account for individual differences in overall axonal excitability rather than to imply a mechanistic association between rheobase and LAh, which have been shown to be only weakly coupled (Shimatani et al., 2023). Linearity was assessed via scatter and partial regression plots; normality was evaluated using the Shapiro-Wilk test. Outliers were screened using boxplots (univariate) and Mahalanobis distance (multivariate).

All anonymized data and supplementary materials supporting the findings of this study are available in Figshare (Naidoo and Jones, 2025).

## 3. Results

### 3.1 Response Probability

Skin temperature remained stable throughout the experiment (above 32), with a paired mean difference from start to end of 1.15 (95.0% CI 0.67 - 1.77). This confirmed that changes in response probability were not confounded by temperature drift. Representative waveform data from a single participant illustrate the method used to quantify response probability after 10 repeated stimuli of each train length (Fig. 2). As the number of stimuli per train increased, the likelihood of evoking a CMAP in the APB muscle increased. Each condition (defined by a specific pulse width and amplitude) was tested three times to obtain a reliable estimate of the mean response probability for each participant. As summarized in Table 1, response probability increased systematically with both increasing pulse amplitude and longer pulse widths. Notably, only one condition (80% amplitude, with a 0.1 ms pulse width) had a group mean response probability below the 5% criterion used to define the Subthreshold Susceptibility Index (SSI).

**Table 1.** Mean ± SD response probability (%) for each combination of pulse amplitude and pulse width (n = 15). For each participant, response probabilities were averaged across all trains and cycles for each width-amplitude combination; group means ± SD are shown. Only one condition (80% amplitude, 0.1 ms pulse width) yielded a group mean response probability < 5%, indicating a low likelihood of inadvertent CMAP generation during subthreshold stimulation with these parameters.

| Pulse Amplitude (% of threshold) | 0.1 ms | 0.5 ms | 1.0 ms |
| --- | --- | --- | --- |
| 80 | 1.6 ± 6.6 % | 11.4 ± 23.6 % | 37.0 ± 41.8 % |
| 85 | 9.6 ± 22.4 % | 34.7 ± 40.2 % | 55.7 ± 45.0 % |
| 90 | 24.9 ± 35.8 % | 55.8 ± 44.3 % | 64.6 ± 44.7 % |

A two-way repeated measures ANOVA revealed a significant interaction between amplitude and width on response probability (F(4,56) = 4.853, p = .002, partial η² = .257). Follow-up analysis of simple main effects showed that amplitude significantly influenced response probability at each width (Fig. 3). At 80% of threshold, pulse width had a significant effect on the response probability, F(2,28) = 29.399, p < .001, with all pairwise comparisons reaching significance (p ≤ .015). At 85% and 90% of threshold, the effects of pulse width were also significant, F(2, 28) = 43.881, p < .001 and F(2, 28) = 34.243, p < .001, with all pairwise comparisons statistically significant (p ≤ .002 & p ≤ .023, respectively). Overall, the pattern of results indicates that both greater amplitude and longer pulse width increase the likelihood of evoking a CMAP, with all pairwise comparisons across conditions showing statistical significance (p ≤ .023; Fig. 3).

**Fig 3.**
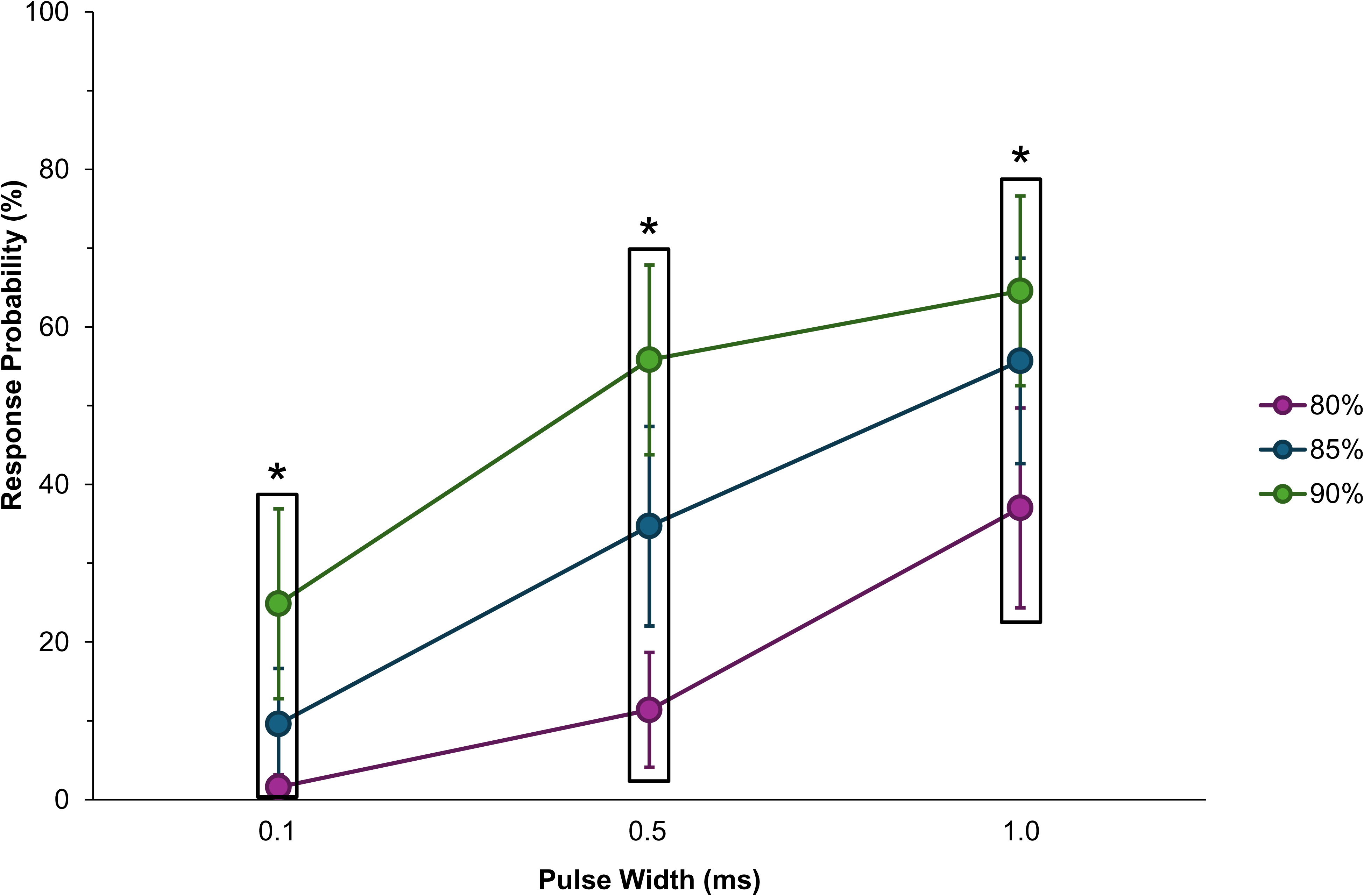
Mean probability of a CMAP response (± 95% CI) as a function of pulse width for three amplitudes (% of motor threshold). Response probability increased with both amplitude and pulse width, with a significant amplitude x width interaction. For clarity, pulse width is shown on the x-axis; an alternative arrangement using amplitude on the x-axis was considered but deemed redundant, as statistical inferences were consistent across both dimensions (all pairwise comparisons p ≤ .023). Boxed groups denote the three amplitudes at each pulse width, and an asterisk (*) above each box indicates that all pairwise comparisons within that pulse width were statistically significant.

The results above report mean response probability averaged across train length and therefore do not reveal how response probability evolves with additional pulses. To address this, we examined the relationship between the number of pulses per train and the likelihood of evoking a CMAP for each amplitude-width combination. Response probability for each condition was plotted as a function of pulse number and fit with a sigmoid function (Equation 1). Figure 4 shows these response curves, separated by pulse width (Panels A–C) and colour-coded by amplitude. Each point represents the median of the 15 participants’ mean response probabilities. At each pulse width, higher amplitudes produced steeper curves, indicating that fewer pulses were needed to reach high response probabilities.

**Fig 4.**
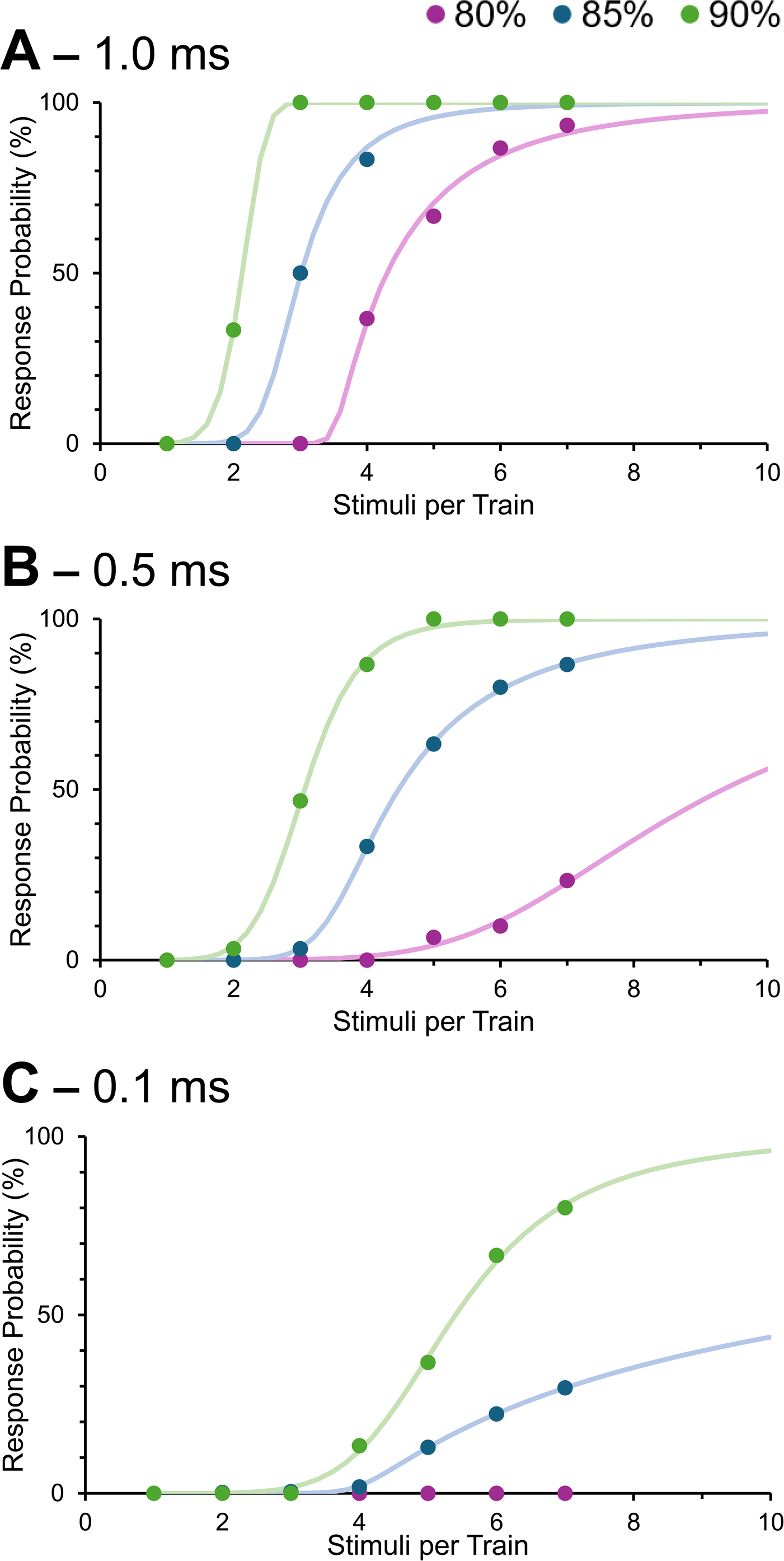
Response probability (%) as a function of the number of pulses per train for pulse widths of (A) 1.0 ms, (B) 0.5 ms, and (C) 0.1 ms. Each data point represents the median response probability across participants for a given train length. Curves were fitted to the data using an asymmetric sigmoid function (Eq. 1) for each combination of pulse width and subthreshold amplitude. The grey dashed line indicates the 5% response probability criterion used to calculate the Subthreshold Susceptibility Index (SSI). This provides a practical tool for identifying amplitude-width-&-train length combinations that minimize the risk of peripheral activation during IONM.

To summarize these effects, we computed the Subthreshold Susceptibility Index (SSI), defined as the number of stimuli required to reach a 5% probability of eliciting a CMAP. This threshold is indicated by the intersection of each fitted curve with the horizontal dotted grey line (Fig. 4), and the resulting SSI values are reported in Table 2. Lower SSI values indicate *greater* susceptibility to subthreshold stimulation, i.e. fewer stimuli are needed to reach the 5% response probability criterion. SSI values decreased systematically with increasing pulse amplitude and pulse width, reflecting the additive effects of these parameters on temporal summation. For example, at the highest-intensity condition (90% threshold, 1.0 ms pulse width), fewer than 2 stimuli were required to reach the 5% threshold. In contrast, at lower intensities (e.g. 80% and 0.5 ms), about 5 stimuli were required. For the lowest-intensity condition (80% and 0.1 ms), CMAP responses were too infrequent to permit a stable curve fit, and no SSI value could be estimated.

**Table 2.** Subthreshold Susceptibility Index (SSI) values for each combination of amplitude and width (n = 15). SSI represents the number of stimuli required to reach a 5% probability of evoking a CMAP, derived from the fitted sigmoid function (Eq. 2). No value was obtained for the 80% amplitude, 0.1 ms pulse width condition because responses were insufficient for reliable curve fitting.

| Pulse Amplitude (% of threshold) | 0.1 ms | 0.5 ms | 1.0 ms |
| --- | --- | --- | --- |
| 80 | N/A | 5.1 | 3.5 |
| 85 | 4.3 | 3.1 | 2.3 |
| 90 | 3.5 | 2.1 | 1.6 |

### 3.2 Latent Addition

A moderate positive association was observed between mean response probability and LAh (r(13) = 0.475, p = 0.073; Fig. 5). The regression line illustrates that participants with larger persistent sodium currents tended to exhibit greater susceptibility to subthreshold activation. Each data point in Figure 5 represents a participant’s grand-average response probability, averaged across all pulse widths and amplitudes.

**Fig 5.**
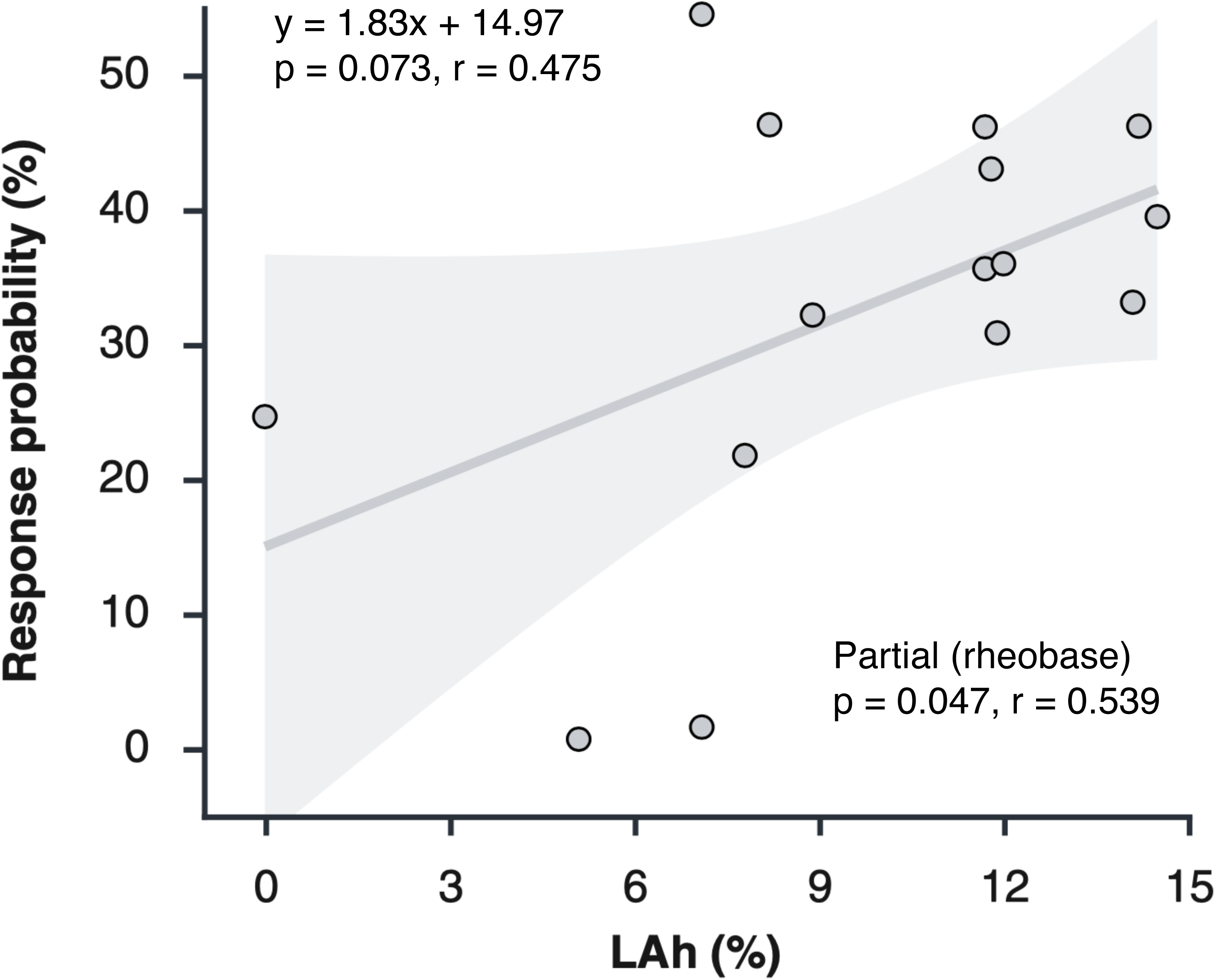
Relationship between mean CMAP response probability and LAh. Each point represents one participant (n = 15). A positive association was observed between Lah and response probability (regression line), which strengthened after controlling for rheobase (r_partial_= .539, p = .047). LAh accounted for approximately 22.5% of the variance in response probability in the bivariate analysis and 29% after adjustment for rheobase, suggesting that stronger persistent sodium current is linked to greater susceptibility to subthreshold activation, although additional factors contribute to the overall variability.

To examine whether this relationship was influenced by individual differences in baseline excitability or passive membrane properties, partial correlations were performed using rheobase and the fast time constant (τ_1_) as covariates. Controlling for τ_1_ slightly reduced the correlation (r_partial_ = 0.404), indicating that variability in τ_1_ did not influence the association between LAh and response probability. In contrast, controlling for rheobase strengthened the correlation (r_partial_(12) = 0.539, p = 0.047), suggesting that the variability in baseline excitability partially masked the association between LAh and response probability. LAh accounted for approximately 22.5% of the variance in response probability in the bivariate analysis and 29.0% after adjusting for rheobase, indicating that persistent sodium current explains a modest but meaningful proportion of inter-individual variability in subthreshold responsiveness.

## 4. Discussion

This study demonstrated that the likelihood of evoking a compound muscle action potential (CMAP) in response to trains of subthreshold stimuli increases with both greater stimulus amplitude and longer pulse width, and that these two parameters interact in a nonlinear manner to modulate excitability. Importantly, individuals with greater susceptibility to this form of temporal summation, reflected by higher response probabilities and lower Subthreshold Susceptibility Index (SSI) values, also exhibited larger persistent sodium currents, as estimated by the LAh metric. This represents the first empirical association between the subthreshold superexcitability phenomenon and an established biophysical marker of persistent inward sodium conductance, providing a partial mechanistic explanation for inter-individual variability in susceptibility to peripheral nerve activation by subthreshold stimulus trains.

### 4.1. Practical Implications for Intraoperative Neuromonitoring

These findings have direct implications for optimizing intraoperative stimulation parameters, particularly in balancing safety margins with reliable CMAP detection. They also extend the foundational work by Urriza et al. (2016), who first described CMAP generation from trains of subthreshold stimuli using a single stimulation condition (90% of threshold amplitude, 0.5 ms pulse width). That study demonstrated the potential for peripheral nerve activation without transsynaptic activation of corticobulbar pathways, raising concerns for intraoperative neuromonitoring (IONM). Our factorial design addressed unanswered questions by systematically varying amplitude and pulse width to determine how these parameters interact to influence response probability, quantified using mean response probability and the SSI.

We, somewhat arbitrarily, defined a clinically meaningful activation risk as a 5% response probability and calculated SSI values to determine how many stimuli for each condition could be safely used before reaching this threshold. Without any limit on train length, only the lowest stimulation condition tested (80% amplitude, 0.1 ms) met this safety threshold for trains up to seven stimuli; in this case, SSI could not be estimated because responses never reached 5%. All other conditions crossed the 5% threshold with fewer than seven pulses, showing that modest changes in stimulus parameters can substantially alter subthreshold superexcitability.

From a practical standpoint, IONM practitioners may prefer to limit train length rather than rely on a single ‘*safe*’ amplitude-width setting. Table 2 provides a framework: if the upper limit is set to four pulses per train, as originally suggested by Dong et al. (2005), any condition with an SSI ≥ 4.0 can be considered acceptable. Under this criterion, three combinations may be used: 80% with 0.1 ms, 80% with 0.5 ms, and 85% with 0.1 ms, giving greater flexibility. With a limit of three pulses, an even broader set of combinations qualifies, allowing for individualized trade-offs between risk and successful monitoring.

Although these parameters are based on median nerve data, similar principles may apply to cranial motor nerves if their susceptibility to subthreshold superexcitability proves comparable to that observed here. Confirmatory studies in the facial nerve are needed to establish whether these recommendations generalize.

These findings also highlight inconsistencies in current practice. Protocols for corticobulbar tract monitoring vary widely: 3–5 pulses, pulse widths from 0.05 to 0.5 ms, and interstimulus intervals of 1–2 ms, yet rarely specify amplitude as a percentage of single-pulse motor threshold (Izzo et al., 2024; Verst et al., 2012; Dong et al., 2005). This omission is notable, given that amplitude is a key determinant of susceptibility and was central to the original warning that threshold in peripheral nerves is not fixed and that subthreshold superexcitability needs to be considered when interpreting IONM responses (Téllez et al., 2016). Although Tellez et al. (2016) recommended that CoMEP settings should be 10-30% below threshold, our data shows that this is a broad range, and that 10% below threshold is drastically different than a subthreshold pulse that is 20% below threshold. The lack of standardized, threshold-referenced amplitude reporting hampers reproducibility and may contribute to variability in false-negative rates across centers.

### 4.2. A Partial Mechanistic Explanation - Persistent Sodium Currents

While the results above provide IONM practitioners with practical options for minimizing risk, they do not explain why some individuals remain more susceptible to subthreshold activation than others. Inter-individual variability was anticipated from the outset, but its mechanistic basis has remained largely speculative in the literature (Téllez et al., 2016; Urriza et al., 2016; Urriza, 2020). To address this, we incorporated the latent addition (LAh) metric (Bostock and Rothwell, 1997) as an empirical indicator of persistent sodium conductance in axons.

A moderate correlation between LAh and response probability suggests that persistent sodium current contributes to, but does not fully account for, susceptibility to subthreshold superexcitability. These currents provide a sustained inward sodium conductance that does not inactivate rapidly, allowing small depolarizations to summate over successive subthreshold stimuli (Crill, 1996; Baker and Bostock, 1997; Goldin, 2003). We had anticipated a stronger association. LAh explained about 22.5% of the variance in response probability, increasing to 29% after adjusting for rheobase, indicating a modest but measurable contribution.

Recent work by Shimatani et al. (2023) showed that LAh is largely stable across CMAP targets from 10% to 60% Mmax, supporting the validity of our 40% target and suggesting that this measure is relatively independent of response amplitude. Their findings underscore that related metrics such as SDTC integrate multiple conductances (Na_p_, K^+^, and HCN) and cannot isolate a single mechanism. Further investigation using approaches that can separate these channel contributions will be required to clarify the basis of inter-individual differences in subthreshold excitability.

### 4.3. Limitations

This study examined the biophysics of subthreshold superexcitability using the median nerve as a model system. Although convenient for controlled testing, these findings cannot be directly generalized to the facial nerve. Eviston and Krishnan (2016) reported that motor axons of the facial nerve exhibit a less pronounced superexcitability compared with the median nerve, likely reflecting factors such as internodal capacitance or more fast K+ (Kv1 family) channels in the juxtaparanodal region. In contrast, rheobase and SDTC values are remarkably similar between facial and median nerves, suggesting that basic membrane properties are largely conserved. These differences highlight the importance of reproducing the present findings in the facial nerve.

Corticobulbar MEP monitoring presents additional challenges related to nerve depth, electrode configuration, and patient-specific factors such as preoperative nerve function and surgical access. Electrode type and placement can alter current density at the nerve, meaning that pulse width and amplitude alone may not fully standardize stimulation parameters across individuals.

The 5% criterion was chosen pragmatically to represent a conservative limit on inadvertent CMAP activation. Determining an acceptable false-negative rate will ultimately require patient-centered and outcome-based evidence.

## 5. Conclusion

This study quantified the parameter dependence of subthreshold superexcitability and identified persistent sodium conductance as a partial mechanistic contributor. By showing that CMAP probability increases with higher amplitude and longer pulse width, and by defining SSI-based criteria for ‘*safe*’ train lengths, we provide IONM practitioners with practical strategies to reduce false-negative responses. At the same time, our results challenge the persistent assumption that a single-pulse threshold defines a fixed level of excitability, underscoring the need for threshold-referenced, standardized stimulation protocols. The mechanistic link to LAh extends previous literature but also highlights that susceptibility is multifactorial, warranting further investigation to improve patient outcomes.

## Glossary

**Compound muscle action potential (CMAP) -** The summed electrical response recorded from a muscle following stimulation of its innervating motor axons.

**Latent Addition -** A nerve excitability test used to estimate persistent sodium currents by assessing changes in threshold following paired stimuli.

**Motor evoked potential (MEP) -** The electrical response measured from a muscle following transcranial stimulation of motor pathways involving both upper and lower motor neurons.

**Rheobase -** The slope of the charge–duration relationship obtained from the strength–duration test during the latent addition protocol.

**Subthreshold stimulation -** Stimulation at an intensity below the threshold required to elicit a measurable CMAP.

**Subthreshold superexcitability -** A period of enhanced axonal excitability following a subthreshold stimulus during which the threshold for activation is transiently reduced.

**Subthreshold superexcitability index (SSI) -** The number of pulses in a train required to elicit a 5% probability of a CMAP, used as an operational measure of subthreshold superexcitability.

## Data availability statement

Anonymized data and supporting documents for this study are openly available in Figshare at https://doi.org/10.6084/m9.figshare.29897132.

## Declaration of generative AI and AI-assisted technologies in the writing process

During the preparation of this manuscript, the authors used ChatGPT (OpenAI, San Francisco, CA, USA) to assist with improving clarity, organization, and grammar in selected sections. All AI-generated suggestions were reviewed, edited, and approved by the authors, who take full responsibility for the content of the final manuscript.

## Author Contributions

AN: data acquisition; analysis; visualization; writing (original draft)

KEJ: conceptualization; supervision; statistical analysis; interpretation; writing (review & editing)

All authors approved the final version of the manuscript and agree to be accountable for all aspects of the work.

## Acknowledgements

We would like to thank Dr. Siyu Du, whose work as a clinical research coordinator facilitated various aspects of the study. We would also like to extend our gratitude to Dr. James Howells, who facilitated the improvement of QTracS, allowing us to obtain images for this paper. Lastly, we would like to express our appreciation to all the participants who took part in this study.

## Abbreviations

APB: Abductor pollicis brevis
CMAP: Compound muscle action potential
EMG: Electromyography
IONM: Intraoperative neuromonitoring
LAh: Latent addition induced change in threshold (at a 0.2 ms delay)
MEP: Motor-evoked potential
SSI: Subthreshold susceptibility index
TES: Transcranial electrical stimulation

